# Analysis of cutoff point estimation for determining seropositivity in the context of SARS-CoV-2 infections

**DOI:** 10.1101/2021.12.20.21268100

**Authors:** Tiago Dias Domingues, Helena Mouriño, Nuno Sepúlveda

## Abstract

In this work will apply mixture models based on distributions from the SMSN family to antibody data against four SARS-CoV-2 virus antigens. Furthermore, since the true infection status of individuals is known *a priori*, performance measures will be calculated for the methods proposed for cutoff point estimation such as sensitivity, specificity and accuracy. The results of a simulation study will also be presented.

## 1 Introduction

Severe acute respiratory syndrome coronavirus 2 (SARS-CoV-2) infection that causes the devastating and often lethal COVID-19 disease was first detected in China, province of Wuhan in December 2019 ([26]). Rapidly, SARS-CoV-2 infection spread over the entire world and the COVID-19 disease was declared as a pandemic by the World Health Organization.

The detection of the virus is so far done by the so-called reverse transcription quantitative PCR (RT-qPCR) on samples from nasopharyngeal or throat swabs ([26]). In general, only symptomatic individuals or people who were in close contact with detected cases are tested, which might lead to an underestimation of the proportion of individuals infected with SARS-CoV-2 ([31]). Alternatively, serological testing allows to detect asymptomatic individuals exposed to the infection. In addition, serological testing is able to quantify the degree of exposure to the infection in the population. In this context, it is important to estimate seroprevalence at the population level, i.e., the proportion of seropositive individuals that show antibodies against any SARS-CoV-2 antigen ([14]).

The presence of antibodies in a serum sample can be regarded as an indicator of immunity against a given infectious agent or as an indicator of past infection in the absence of vaccination ([10]). The detection of antibodies in the serum samples is classically done via enzyme linked immunosorbent assays (ELISA), where the resulting data are light intensities, also called optical density, which reflects the underlying antibody concentration in the samples ([9]). For statistical convenience, the analysis of serological data proceeds by dichotomizing the amount of antibodies present in the serum of an individual using an arbitrary cutoff point in the antibody distribution to achieve a certain sensitivity and specificity. This allows the classification of individuals into seronegative (with antibody levels below the cutoff point) and seropositive (with antibody levels above the cutoff point) ([26]).

Given the possible impact of the cutoff chosen, different criteria for seropositivity determination have a direct impact on the sensitivity and specificity of the respective serological classification ([22]). In addition, it might also impact the estimation of the seroprevalence ([13]) and the following (epidemiological) decision that can be taken when facing a given estimate of this epidemiological parameter. This means that when determining the cutoff point for a serological test, one should take into account the benefit of the test, the economic and social consequences of serological misclassification and the prevalence of the disease in the population. It turns out that these aspects are often ignored in practice ([25]).

One of the traditional methods to establish the cutoff point in serological assays is to consider the logarithmic transformation of the antibody concentration of a known seronegative population and proceed to calculate the mean plus 2 or 3 standard deviations ([4,18,25,32]). This method is more adequate when the antibody distribution of the seronegative population is normally distributed ([4]). However, our previous studies of different serological data ([9,30]) showed evidence against a normality assumption for the antibody levels associated with a putative seronegative population. In the case where the true infection (or disease) status is known, ROC curve-based methods are most commonly used to determine the cutoff point for defining seropositivity. These methods are widely discussed in the literature ([5,11,12,21,23,27,33]).

Alternatively, finite mixture models can be used to determine the seropositivite cutoff directly from the data ([4,9,13,21,29]). In our previous work, three methods for determining seropositivity cutoff were explored using the so-called scale mixtures of Skew-normal distributions in the case where the true infection status is unknown ([9]). In this paper we applied the same methods and models in order to evaluate their performance in freely available serological data concerning SARS-CoV-2 virus ([26]). We also used simulation to understand the performance of the cutoff estimators associated with different criteria for seropositivity determination.

## 2 Serological data concerning SARS-CoV-2 virus

In this study we analyzed IgG antibody responses against four SARS-CoV-2 spike or nucleoprotein antigens: RBD – glycoprotein receptor-binding domain; *S*^*tri*^ — S trimeric spike protein; S1 — spike glycoprotein S1 domain; S2 – SARS-CoV-2 spike glycoprotein S2 domain. Antibodies were measured in serum samples collected up to 39 days after symptom onset from 215 adults in four French hospitals (53 patients and 162 health-care workers) with quantitative RT-PCR-confirmed SARS-CoV-2 infection. A total of 335 negative control serum samples were collected from France, Thailand, and Peru before the start of the COVID-19 pandemic ([26]). A detailed description of lab procedures can be found in the original study ([26]). The data is freely available at https://github.com/MWhite-InstitutPasteur/SARSCoV2SeroDXphase2.

## 3 Statistical methods

Serological data can be viewed as arising from two or more latent populations; each population is assumed to represent different levels of exposure to a given antigen. For simplicity, individuals that were never exposed or exposed a long time ago to an infectious agent are considered as seronegative. In contrast, individuals exposed to the same infectious agent are considered seropositive. In this scenario, the antibody distribution can be described by a mixture of two or more probability distributions ([8]). However, the true serological state of the individuals is unknown and therefore it needs to be estimated.

In the particular case of the SARS-CoV-2 data, we know which individuals were exposed to the virus and, therefore, we can assume to know which individuals are true seronegative and seropositives.

In many serological studies, it is common to assume a normal distribution for the basis of the mixture models. However, the behaviour of antibody distribution is not constant over time and their concentration decreases after infection ([26]). This fact makes the distribution of the seropositive population skewed to the left ([10]). In order to accommodate the possible skewness in the seropositive population we use the scale mixture of Skew-Normal (SMSN) class of distributions that include the Skew-Normal and the Skew-t distributions, which will be the focus of our study. A brief description of these alternative distributions can be found below.

### 3.1 Skew-Normal and Skew-t distributions

Let *W* ⌢ *SN*(*µ, σ*^2^, *α*) a random variable with a Skew-Normal distribution. In this distribution, the parameters *µ, σ*^2^, and *α* can be seen as the location, scale, and shape parameters, respectively. Then the probability density function (pdf) is given by

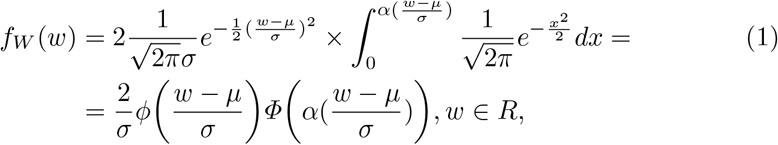

where *ϕ*(·) and *Φ*(·) is the pdf and the cumulative distribution function of the standard Normal distribution, respectively ([1,3,9]). The Skew-Normal distribution is part of a family of distributions called the Scale Mixtures of Skew-Normal distributions (SMSN), of which the Skew-t distribution is also a particular case ([9]).

A random variable *W* is said to have a Skew-t distribution, *W* ⌢ *ST* (*µ, σ*^2^, *α, v*), if the pdf is given by

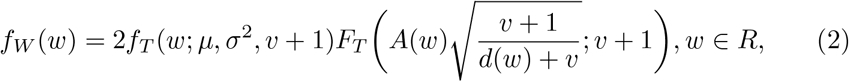

where *f*_*T*_ (.; *µ, σ*^2^, *v* + 1) and *F*_*T*_ (.; *µ, σ*^2^, *v* + 1) represents the pdf and the cumulative distribution function of the generalized Student’s t distribution with *v* + 1 degress of freedom, 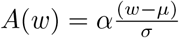 and 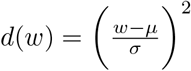 ([1,3,9]).

### 3.2 Finite mixture models

Let *G*_1_ and *G*_2_ be the seronegative and seropositive subpopulations from a population *G*, respectively. Let π_1_ and π_2_ the probabilities of sampling a seronegative and a seropositive individual, respectively (with the usual restriction of 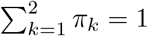 and 0 ≤ *π*_*k*_ ≤ 1) and considering *Z* the random variable that represents the antibody level. The probability density function (pdf) of *Z* is given by

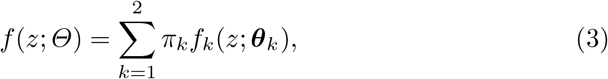

where *f*_*k*_(*z*; ***θ***_*k*_) is the mixing probability density function of *Z* associated with the *k* − *th* latent population and parameterized by the vector ***θ***_*k*_. *Θ* is the vector of all unknown parameters of the mixture model, i.e., *Θ* = (*π*_1_, *π*_2_, ***θ***_1_, ***θ***_2_). In our application, *f*_*k*_(*z*; ***θ***_*k*_), is given by the Skew-normal or the Skew-t distributions.

In general, the estimation of a finite mixture model can be done by the classical EM algorithm ([15]). The EM algorithm is an iterative method widely used in incomplete data problems where the maximum likelihood estimators (MLE) have no closed expression ([7]). Considering (*z*_1_, *z*_2_, …, *z*_*n*_) the observed sample of size *n* and *Y*_*i*_ ≡ *Y*_*ik*_, (*i* = 1,.., *n*; *k* = 1, 2), the binary vector representing the component from which the data comes from. Thus, *Y*_*i*_ ⌢ *Bernoulli*(*π*_2_) and the pdf of *Y*_*i*_ is given by

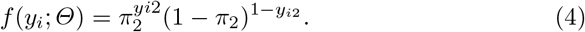

We have that the complete data is the pair (*z*_*n*_, *y*_*n*_) and the joint pdf is given by

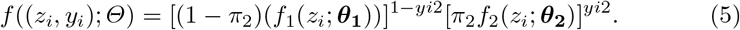

Then, the log-likelihood function is given by

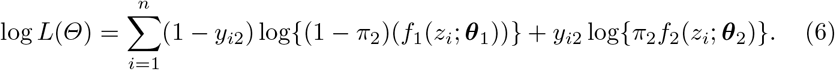

The step E of the EM algorithm consists in obtaining

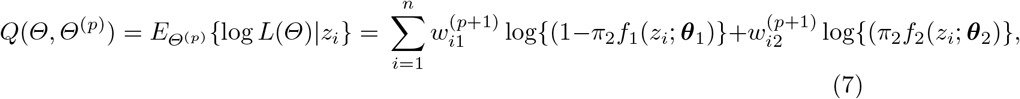

where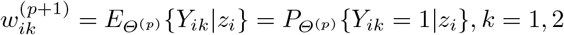

The step M consists in maximizing *Q*(*Θ, Θ*^(*p*)^) as function of the unknown parameters. However, if the model has many parameters that need to be estimated, then step M may incur in computational problems such as excessive time consuming or estimate instability. In this sense, it is possible to break the step M into several sub-steps (*S* > 1) that allow to get around these computational constraints by performing some restrictions on the parameters. This method is called expectation-conditional-maximization (ECM) algorithm ([19,17,20]). Considering that *Θ*^(*p*+*s*)^ represents the value of *Θ* in the s^*th*^ CM step of the iteration *p* + 1 in order to maximize *Q*(*Θ, Θ*^(*p*)^) and the constraint function *g*_*s*_(*Θ*) = *g*_*s*_(*Θ*^(*p*+(*s−*1)^), the ECM algorithm is performed as follow ([17]):

1. calculate the expected complete-data log-likelihood given the current estimates of the parameters, *Θ*^(*p*)^. The calculations are the same as for the EM algorithm;
2. fix *Θ*^(*p*)^ and calculate *Θ*^(*p*+*s*)^ to maximise the expected complete-data loglikelihood;
3. fix *Θ*^(*p*+*s*)^ and calculate *Θ*^(*p*+(*s*+1))^ to maximise the expected complete-data log-likelihood on the *s* + 1 sub-step iteration and continuing until you have gone through all the *S* sub-steps.

In this way, it can be seen that *Q*(*Θ, Θ*^(*p*+*s*)^ ≥ *Q*(*Θ, Θ*^(*p*)^) for all *Θ* ∈ *Ω*_*s*_(*Θ*^(*p*+*s*)^), where *Ω*_*s*_(*Θ*^(*p*+*s*)^) = {*Θ* ∈ *Ω* : *g*_*s*_(*Θ*) = *g*_*s*_(*Θ*^(*p*+(*s−*1)^)} ([19,17,20]).

Considering the SMSN family of distributions, namely the Skew-Normal and the Skew-t distributions, the application of the ECM algorithm in the context of mixtures can be found in ([3,16]).

In order to decide which model is the best one among all the models fitted to the same data, we used the Bayesian Information Criterion (BIC) ([9]).

### 3.3 Definition of seropositivity

Seroprevalence is an epidemiological measure defined by the proportion of seropositive individuals in the sample. For its estimation, it is then necessary to define the serological status of the *i*-th individual by dychotomization the variable, *Z*_*i*_, which represents the antibody concentration of the individual. This dychotomization is done by determining a value *c* such that for antibody values equal to or greater than *c*, the individual is classified as seropositive and seronegative, otherwise. Thus, let *Y* be the random variable representing the number of seropositive individuals in a sample of size *n*, we have to

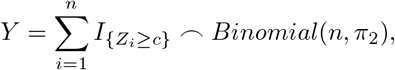

where *π*_2_ represents the seroprevalence, i.e, *π*_2_ = *P*[*Z*_*i*_ ≥ *c*] and *I*_{*·*}_ is the indicator variable. Considering that the random variable representing the antibody levels *Z*_*i*_ is modelled by a finite mixture of distributions, the way to estimate the cutoff *c* from the observed data is not standard. To facilitate the determination of this cutoff value, we below present three estimation methods or criteria.

- **Method 1 (M1):** It is based on the 99.9%-quantile associated with the estimated seronegative population. This method is the most popular in seroepidemiology ([28,29]). It is often called as the 3*σ* rule, because the 99.9%-quantile is given by the mean plus 3 times the standard deviation of a normally distributed seronegative population;
- **Method 2 (M2):** It relies on the minimum of the density mixture functions. In the case of two latent populations, the cutoff corresponds to the absolute minimum, and in the case of three or more latent populations the cutoff corresponds to the lowest relative minimum. This point can be calculated using the Dekker’s algorithm ([6]). It should be noted that the minimum of the mixing function is not expected to coincide with the point of intersection of the probability densities of each individual subpopulation;
- **Method 3 (M3):** It imposes a threshold in the the so-called conditional classification curves ([29]). Under the assumption that all components but the first one refer to seropositive individuals, the conditional classification curve for the *i*-th individual given the antibody level *Z*_*i*_ = *x* is defined as

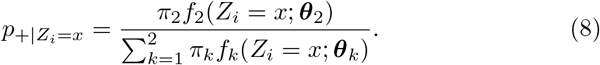

In turn, the classification curve of seronegative individuals is simply given by

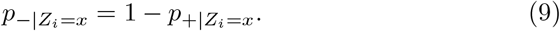

After calculating these curves, one can impose a minimum value for the classification of each individual. In this case, two cutoff values arise in the antibody distribution, one for the seronegative individuals and another for seropositive individuals. Mathematically, the classification rule is given as follows

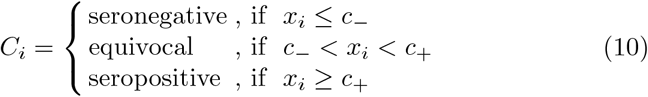

where *c*_*−*_ and *c*_+_ are the cutoff values in the antibody distribution that ensure a minimum classification probability, say 90%. To calculate these cutoff values in practice, one can use the bisection method providing an initial interval where they might be located ([29]).

### 3.4 Performance of the proposed methods for cutoff point estimation

In order to evaluate the performance of each of the cutoff points, we estimated the respective sensitivity and specificity. Let *D* and *D*^*∗*^ be the true and estimated serological classification (or infection status), respectively. Sensitivity is defined as the conditional probability

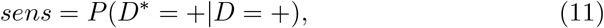

In turn, the specificity i s defined as

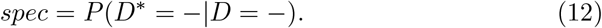

The overall performance of each method is given by the accuracy (ACC) of the proposed method which corresponds to the proportion of correct results, that is,

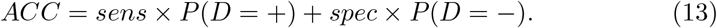

### 3.5 Simulation study

We performed a small simulation study to assess the performance of cutoff points proposed by each method. With this purpose, we assume two simulation scenarios regarding the mixture model assumed for the data: (i) a mixture model based on the Skew-Normal distributions and (ii) a mixture model based on the Skew-t distribution.

For each scenario, we simulated 1000 samples with dimensions 100, 500 and 1000. In addition, for each simulation cycle, the weight of the mixture model was varied to check the ability of the model to identify the seropositive component even when the weight assigned to that component is very low. The implications of varying the weight of the seronegative and seropositive population are as follows: in the case where the proportion of seronegative individuals is very high relative to seropositive individuals, more effective decisions can be made to control the number of infections in the population. The opposite scenario is important in the case of effectiveness of vaccination in the population, particularly for individuals who may have lost immunity.

To this end, it was considered that the proportion of seronegative individuals could take the values 90%, 60% and 30%, being the respective proportion of seropositive individuals 10%, 40% and 70%, respectively. For each simulated sample, the parameters of the mixture model were estimated by maximum likelihood (via the EM algorithm) according to the distributional scenarios described above, as well as the respective cutoff points according to the methods M1, M2 and M3. Considering *θ*^*∗*^ the estimated parameter, *θ* the true value of the parameter, than we calculate the relative error that is 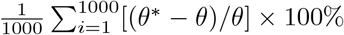 and the mean squared error (MSE), i.e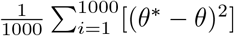.

### 3.6 R packages

We used the package `mixsmsn` to fit different mixture models based on SMSN ([24]). In particular, we used the function `smsn.mix` to estimate the model parameter via the EM algorithm For fitting the Student’s t-distribution, we considered the R package `extraDistr` ([34]), namely, the function `dlst` to calculate their density, the function `plst` to define the cumulative distribution function and the function `rlst` to generate random samples in the simulation study. The fitting of the Skew-Normal distributions was performed with the package `sn` ([2]). The functions `dsn`, `psn` and `rsn` were used to calculate the probability density function, the cumulative distribution function and generate random samples of the Skew-Normal distribution, respectively. In the case of the Skew-t distribution, the functions `dst`, `pst` and `rst` were used to calculate the probability density function, the cumulative distribution function and generate random samples, respectively.

## 4 Results

### 4.1 Patients characteristic’s

For this study, data relating to 549 individuals was analysed. Serum samples were collected from individuals with confirmed SARS-CoV-2 infection by PCR test in four hospital units from Paris, namely: 4 (0.7%) from the Hôpital Bichat, 49 (9.0%) from the Hôpital Cochin and 161 (29.3%) from the Nouvel Hôpital (Strasbourg). Regarding the negative controls, 68 (12.4%) are from the Thai Red Cross (TRC), 90 (16.4%) from the Peruvian donors (NHP) and 177 (32.2%) from the France blood donors (Établissement Français du Sang). For each antigen under analysis, the logarithmic transformation of base 10 was considered for the concentration of antibodies against that antigen.

Regarding the analysis of antibodies by the individuals who performed PCR test, there were statistically significant differences between individuals who tested negative and positive for SARS-CoV-2 by Mann-Whitney test (RBD: 1.64 vs. 3.48, *p <* 0.001; S1: 1.72 vs. 2.59, *p <* 0.001; S2: 1.79 vs. 2.99, *p <* 0.001; Stri: 1.59 vs. 3.43, *p <* 0.001) (Figure 1). Such differences were expected given the general knowledge about the infection status, i.e., individuals who have already been exposed to the virus will have a higher concentration of antibodies than those who are still susceptible.

**Fig. 1:**
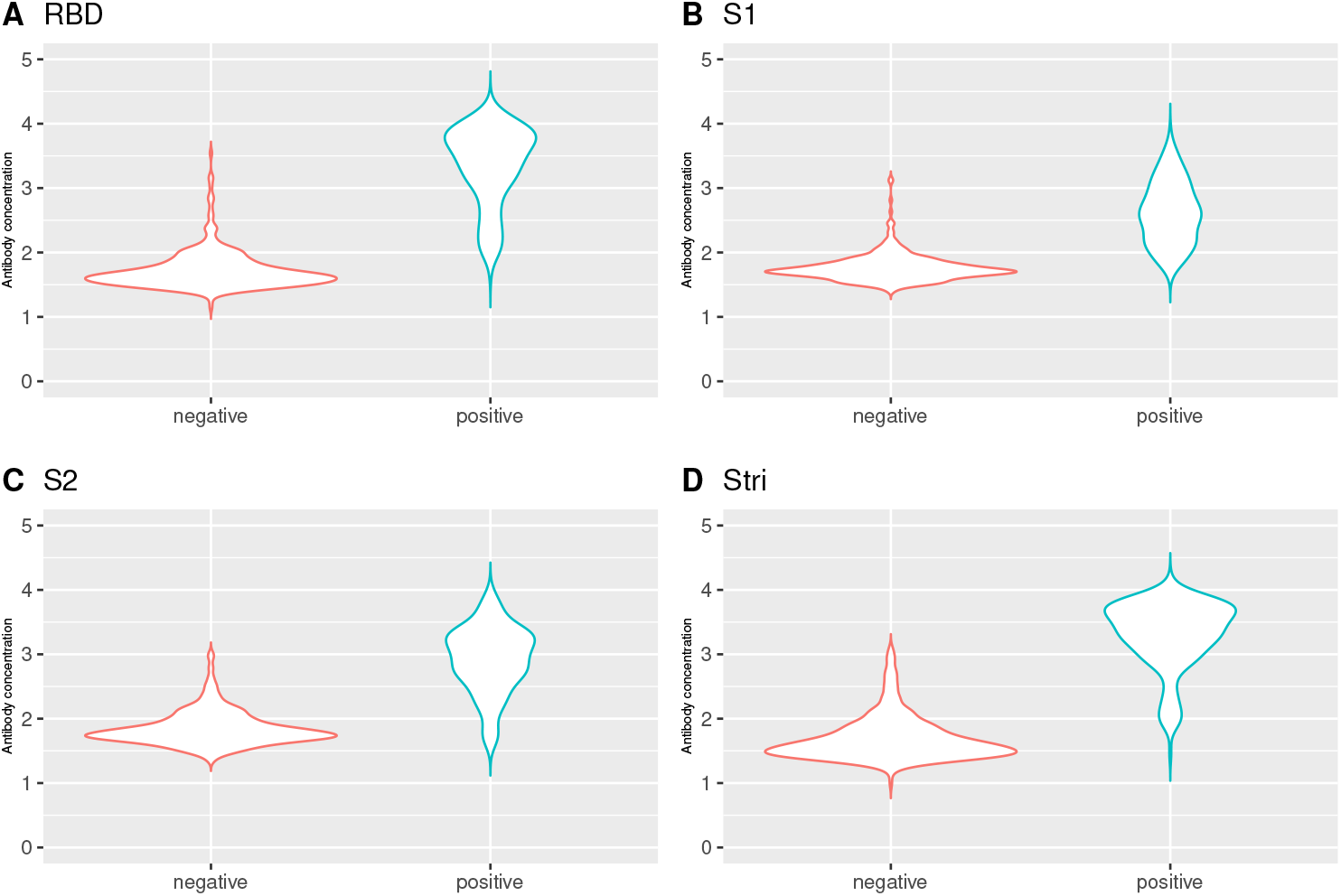
Antibody distribution by infection status. **A**. Antibody distribution for RBD antigen. **B**. Antibody distribution for S1 antigen. **C**. Antibody distribution for S2 antigen. **D**. Antibody distribution for *S*^*tri*^ antigen. Number of negative individuals: 335; number of positive individuals: 214. Antibody concentration in y axis is given in log10 units.

### 4.2 Mixture Model approach

We performed the fitting of the different mixture models considering two subpopulations, i.e., a seronegative population and a seropositive population. According to the BIC values, the model based on the Skew-Normal distribution was considered for the following antigens: RBD (BIC=852.25), S1 (BIC=561.63), S2 (BIC=775.29). For the case of the Stri antigen, the best model was found to be the Skew-t distribution (BIC=915.82) (Figure 2 and table 2). As has been observed in previous studies, there is a marked skew to the right of the data for the seronegative population and a skewed for the left in the seropositive population, although not very marked for the S1 (*α*_*S*1_ = 1.062) and S2 (*α*_*S*2_ = 0.450) antigens (Table 1).

**Fig. 2:**
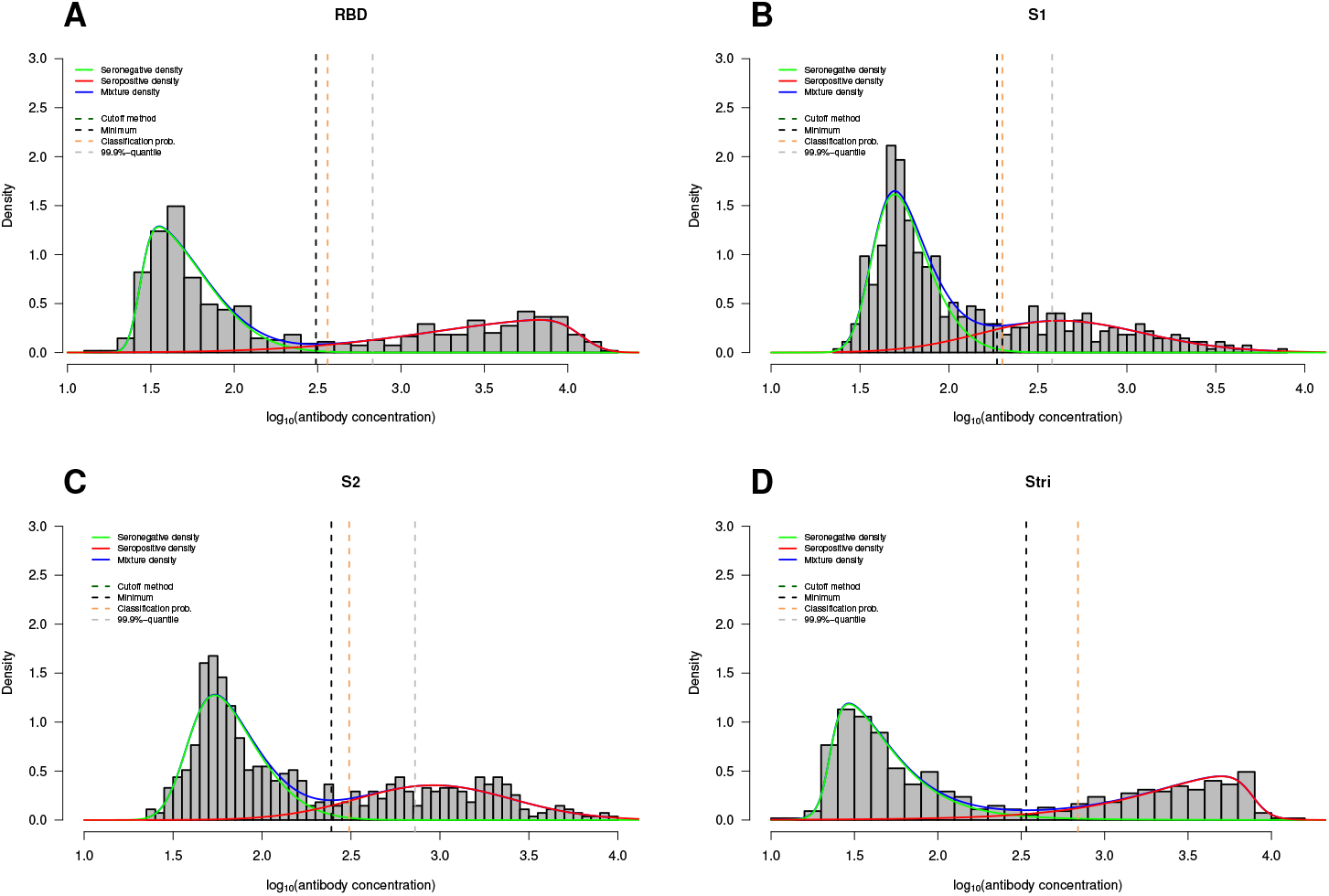
Best models with two components for the data under analysis. **A**. Antibody distribution for RBD antigen. **B**. Antibody distribution for S1 antigen. **C**. Antibody distribution for S2 antigen. **D**. Antibody distribution for *S*^*tri*^ antigen. Antibody concentration in *x* axis is given in log_10_ units.

**Table 1:** Parameter estimates for the best model

| Antigen Distribution |  | Seronegative population |  |  |  | Seropositive population |  |  |  |
| --- | --- | --- | --- | --- | --- | --- | --- | --- | --- |
| | | $\mu$ | $\sigma^2$ | $\alpha$ | $v$ | $\mu$ | $\sigma^2$ | $\alpha$ | $v$ |
| RBD | Skew-Normal | 1.435 | 0.125 | 6.318 | NA | 4.077 | 0.767 | -7.634 | NA |
| S1 | Skew-Normal | 1.569 | 0.062 | 2.687 | NA | 2.339 | 0.321 | 1.062 | NA |
| S2 | Skew-Normal | 1.583 | 0.096 | 2.804 | NA | 2.817 | 0.212 | 0.450 | NA |
| $S^{tri}$ | Skew-t | 1.352 | 0.121 | 5.751 | 4.873 | 3.885 | 0.367 | -6.482 | 4.873 |

**Table 2:**
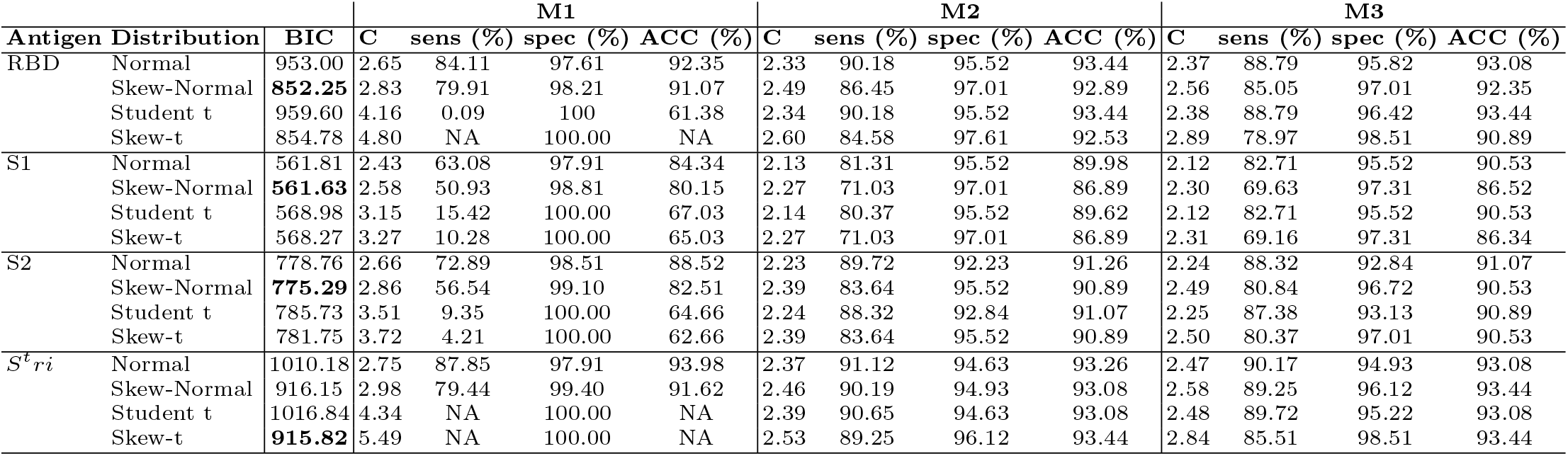
Bayesian Information Criterion (BIC) values, cutoff value estimates, sensitivity, specificity and accuracy for each method by antigen according to the best model. *C* denotes the cutoff point estimate.

### 4.3 Seropositivity estimation

After defining the model that best fits the data, we proceeded to categorize the amount of antibodies for each antigen by estimating the cutoff point. For this, we used the methods M1, M2 and M3 already described and whose results are shown in Figure 1 and table 2.

Estimation of the cutoff point based on the minimum densities of the mixture model (M2) proved to be the method with the highest sensitivity for classifying seropositive individuals, as well as the one that produces the highest proportion of correct results (accuracy) for the RBD antigen (*cutoff* = 2.49, *sens* = 86.45%, *ACC* = 92.89%), S1 (*cutoff* = 2.27, *sens* = 71.03%, *ACC* = 86.89%) and S2 (*cutoff* = 2.39, *sens* = 83.64%, *ACC* = 90.89%). In the case of the S^*tri*^ antigen, it was not possible to calculate the sensitivity and accuracy of the method based on the 99.9%-quantile (M1), given the high values that the quantile assumes leading to the seropositive population being fully absorbed by it. Thus, for comparison purposes, the application of each methods to the Skew-Normal distribution was considered, again verifying that the method based on the minimum densities of the mixture model produces the highest sensitivity (*cutoff* = 2.46, *sens* = 90.19%). However, for this antigen, the method with the highest accuracy is based on the conditional probability (set at 90%) of classifying an individual as being seropositive (ACC=93.44%) (Figure 3 and table 2).

**Fig. 3:**
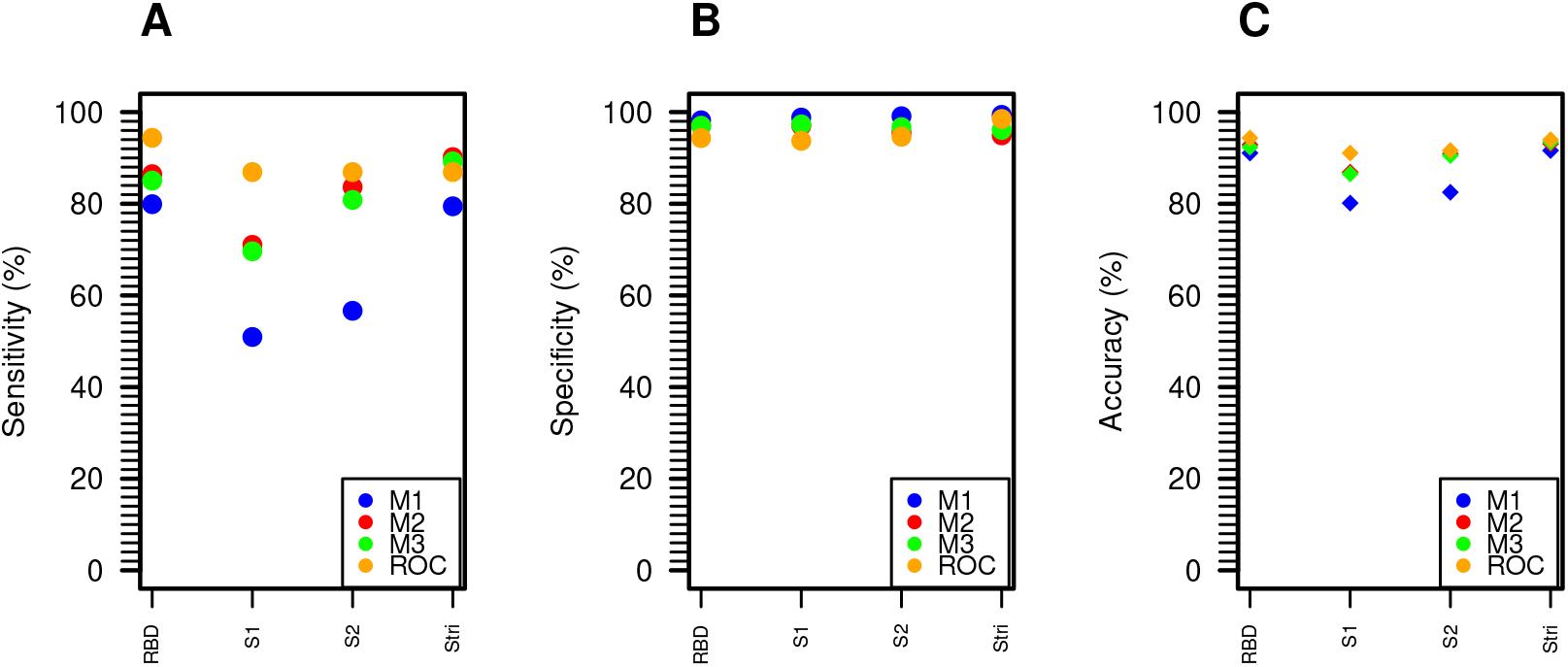
Performance of each method to estimate the cutoff value. **A**. Sensitivity values for each method. **B**. Specificity values for each method. **C**. Accuracy values for each method.

In order to evaluate the quality of methods M1, M2 and M3, the optimal cutoff point was estimated using the ROC curve. This is possible since the true infection status of the individuals is known. It is interesting to see that in terms of specificity and accuracy the results are similar to the method that is traditionally used (ROC curve). However, it is possible to observe a poor performance of the M1 method with regard to its sensitivity. (Figure 3, table 2 and table 3).

**Table 3:** Cutoff point estimates, sensitivity, specificity, accuracy and area under the curve (AUC) for the empirical ROC curve method

| Antigen | Cutoff | Sensitivity (%) | Specificity (%) | Accuracy (%) | AUC (CI 95%) |
| --- | --- | --- | --- | --- | --- |
| RBD | 2.15 | 94.39 | 94.33 | 94.35 | 98.50<br>(97.80, 99.30) |
| S1 | 2.07 | 86.92 | 93.73 | 91.07 | 96.10<br>(94.60, 97.60) |
| S2 | 2.33 | 86.92 | 94.63 | 91.62 | 94.90<br>(92.80, 97.00) |
| $S^{tri}$ | 2.81 | 86.92 | 98.51 | 93.98 | 98.30<br>(97.40, 99.20) |

### 4.4 Simulation results

To conduct the simulation study, two scenarios were considered: the first consists of the scenario where the model that best fits the data is a Skew-Normal distribution, and the second where the model that best fits the data is a Skew-t distribution. For this purpose, the results for the RBD antigen (Skew-Normal distribution) and the S^*tri*^ antigen (Skew-t distribution) were selected. For each scenario the sample size was varied, as well as the proportion of seronegative individuals in the population. The results are shown in table 4 and table 5.

**Table 4:** Relative bias and Mean Square Error (MSE) of the 99.9%-quantile method (M1); minimum of mixture densities method (M2) and conditional probability method (M3) for the RBD antigen. *opt*_*M*1_ denotes the theoretical cutoff point for the 99.9%-quantile; *opt*_*M*2_ denotes the theoretical cutoff point for the minimum of the density mixture method; *opt*_*M*3_ denotes the theoretical cutoff point for conditional probability method. *π*_1_ denotes the weight of the seronegative population; *c*_*M*1_ denotes the cutoff estimated by M1 method after N=1000 simulations; *c*_*M*2_ denotes the cutoff estimated by M2 method after N=1000 simulations; *c*_*M*3_ denotes the cutoff estimated by M3 method after N=1000 simulations.

| **Normal distribution; $opt_{M1} = 2.65$; $opt_{M2} = 2.33$; $opt_{M3} = 2.37$** |  |  |  |  |  |  |  |  |  |  |  |  |
| --- | --- | --- | --- | --- | --- | --- | --- | --- | --- | --- | --- | --- |
| Sample size | $\pi_1$ | $c_{M1}$ | $c_{M2}$ | $c_{M3}$ | Relative bias $c_{M1}$ (%) | MSE ($c_{M1}$) | Relative bias $c_{M2}$ (%) | MSE ($c_{M2}$) | Relative bias $c_{M3}$ (%) | MSE ($c_{M3}$) | % Two comp. retained | |
| 100 | 0.3 | 5.67 | 2.34 | 2.46 | 113.9314 | 0.0914 | 0.6185 | 0.0001 | 3.8938 | 0.0004 | 93.1 |  |
|  | 0.6 | 5.17 | 2.51 | 2.58 | 95.2155 | 0.0641 | 7.6017 | 0.0004 | 8.8272 | 0.0006 | 100.0 |  |
|  | 0.9 | 3.68 | 2.72 | 2.75 | 39.0584 | 0.0114 | 16.6388 | 0.0017 | 15.9472 | 0.0016 | 99.9 |  |
| 500 | 0.3 | 5.68 | 2.35 | 2.48 | 114.3252 | 0.0183 | 0.9958 | 0.000007 | 4.5243 | 0.00003 | 100.0 |  |
|  | 0.6 | 5.19 | 2.51 | 2.59 | 95.9969 | 0.0129 | 7.7858 | 0.00007 | 9.2046 | 0.0001 | 100.0 |  |
|  | 0.9 | 3.72 | 2.70 | 2.73 | 40.3432 | 0.0023 | 16.1593 | 0.0002 | 15.3683 | 0.0002 | 100.0 |  |
| 1000 | 0.3 | 5.69 | 2.35 | 2.47 | 114.6181 | 0.0092 | 0.8462 | 0.000002 | 4.3660 | 0.00001 | 100.0 |  |
|  | 0.6 | 5.19 | 2.51 | 2.59 | 95.9990 | 0.0064 | 7.7956 | 0.00003 | 9.1638 | 0.00005 | 100.0 |  |
|  | 0.9 | 3.73 | 2.70 | 2.73 | 40.6998 | 0.0011 | 16.0422 | 0.0001 | 15.3499 | 0.0001 | 100.0 |  |
| **Skew-Normal distribution; $opt_{M1} = 2.83$; $opt_{M2} = 2.49$; $opt_{M3} = 2.56$** |  |  |  |  |  |  |  |  |  |  |  |  |
| 100 | 0.3 | 4.63 | 2.50 | 2.74 | 63.3181 | 0.0345 | 0.5088 | 0.0001 | 7.0728 | 0.0010 | 96.9 |  |
|  | 0.6 | 5.73 | 2.75 | 2.74 | 102.2463 | 0.0846 | 10.3808 | 0.0010 | 6.9046 | 0.0006 | 99.5 |  |
|  | 0.9 | 3.94 | 3.04 | 2.89 | 39.2453 | 0.0131 | 22.0631 | 0.0043 | 12.7332 | 0.0016 | 94.7 |  |
| 500 | 0.3 | 4.44 | 2.48 | 2.68 | 56.7727 | 0.0053 | -0.5071 | 0.000009 | 4.6945 | 0.00007 | 100.0 |  |
|  | 0.6 | 5.76 | 2.74 | 2.73 | 103.2662 | 0.0171 | 10.2602 | 0.0001 | 6.3299 | 0.00006 | 100.0 |  |
|  | 0.9 | 3.94 | 3.14 | 2.89 | 39.2537 | 0.0025 | 26.1894 | 0.0011 | 13.0415 | 0.0002 | 100.0 |  |
| 1000 | 0.3 | 4.39 | 2.48 | 2.68 | 55.3506 | 0.0024 | -0.5008 | 0.000003 | 4.6036 | 0.00003 | 100.0 |  |
|  | 0.6 | 5.76 | 2.75 | 2.72 | 103.3116 | 0.0085 | 10.4370 | 0.00009 | 6.0958 | 0.00003 | 100.0 |  |
|  | 0.9 | 3.94 | 3.16 | 2.89 | 38.9617 | 0.0012 | 27.1796 | 0.0005 | 12.9545 | 0.0001 | 100.0 |  |
| **Student t distribution; $opt_{M1} = 4.16$; $opt_{M2} = 2.34$; $opt_{M3} = 2.38$** |  |  |  |  |  |  |  |  |  |  |  |  |
| 100 | 0.3 | 5.85 | 2.15 | 2.23 | 40.6111 | 0.0296 | -7.8239 | 0.0004 | -6.3481 | 0.0004 | 99.9 |  |
|  | 0.6 | 15.22 | 2.31 | 2.45 | 265.6374 | 57.9703 | -1.2775 | 0.0001 | 2.7550 | 0.0003 | 100.0 |  |
|  | 0.9 | 33.74 | 2.60 | 2.86 | 710.4484 | 13.8984 | 11.5284 | 0.0013 | 20.084 | 0.0035 | 84.3 |  |
| 500 | 0.3 | 5.85 | 2.16 | 2.25 | 40.4626 | 0.0057 | -7.3517 | 0.00006 | -5.4927 | 0.00004 | 100.0 |  |
|  | 0.6 | 5.39 | 2.31 | 2.47 | 29.3408 | 0.0030 | -0.9376 | 0.00004 | 3.7417 | 0.00006 | 100.0 |  |
|  | 0.9 | 25.38 | 2.59 | 2.92 | 509.6060 | 1.0029 | 11.2388 | 0.0001 | 22.5757 | 0.0006 | 100.0 |  |
| 1000 | 0.3 | 5.85 | 2.16 | 2.25 | 40.5011 | 0.0028 | -7.4162 | 0.00003 | -5.5194 | 0.00002 | 100.0 |  |
|  | 0.6 | 5.37 | 2.31 | 2.47 | 28.8965 | 0.0014 | -1.0348 | 0.000001 | 3.7648 | 0.00001 | 100.0 |  |
|  | 0.9 | 24.52 | 2.59 | 2.93 | 489.0068 | 0.4401 | 11.2467 | 0.00007 | 23.0901 | 0.0003 | 100.0 |  |
| **Skew-t distribution; $opt_{M1} = 4.80$; $opt_{M2} = 2.60$; $opt_{M3} = 2.89$** |  |  |  |  |  |  |  |  |  |  |  |  |
| 100 | 0.3 | 4.59 | 2.35 | 2.53 | -4.5079 | 0.0031 | -9.9120 | 0.0009 | -12.7692 | 0.0021 | 99.3 |  |
|  | 0.6 | 8.14 | 2.47 | 2.68 | 69.4317 | 0.4065 | -5.1031 | 0.0004 | -7.5949 | 0.0012 | 100.0 |  |
|  | 0.9 | NA | 2.78 | 3.06 | NA | NA | 6.6832 | 0.0008 | 5.5875 | 0.0014 | 40.6 |  |
| 500 | 0.3 | 4.43 | 2.31 | 2.49 | -7.8118 | 0.0004 | -11.0747 | 0.0001 | -14.2211 | 0.0003 | 100.0 |  |
|  | 0.6 | 6.81 | 2.48 | 2.71 | 41.8089 | 0.0114 | -4.6537 | 0.00004 | -6.5774 | 0.0001 | 100.0 |  |
|  | 0.9 | 22.93 | 2.83 | 3.22 | 377.5419 | 0.7006 | 8.8679 | 0.0001 | 11.1838 | 0.0005 | 98.7 |  |
| 1000 | 0.3 | 4.42 | 2.32 | 2.50 | -7.9676 | 0.0002 | -10.8456 | 0.00008 | -13.6653 | 0.0001 | 100.0 |  |
|  | 0.6 | 6.82 | 2.49 | 2.73 | 42.1175 | 0.0048 | -4.2254 | 0.00001 | -5.8102 | 0.00004 | 100.0 |  |
|  | 0.9 | 22.81 | 2.85 | 3.28 | 374.9435 | 0.3352 | 9.6048 | 0.00007 | 13.0902 | 0.0002 | 99.5 |  |

**Table 5:**
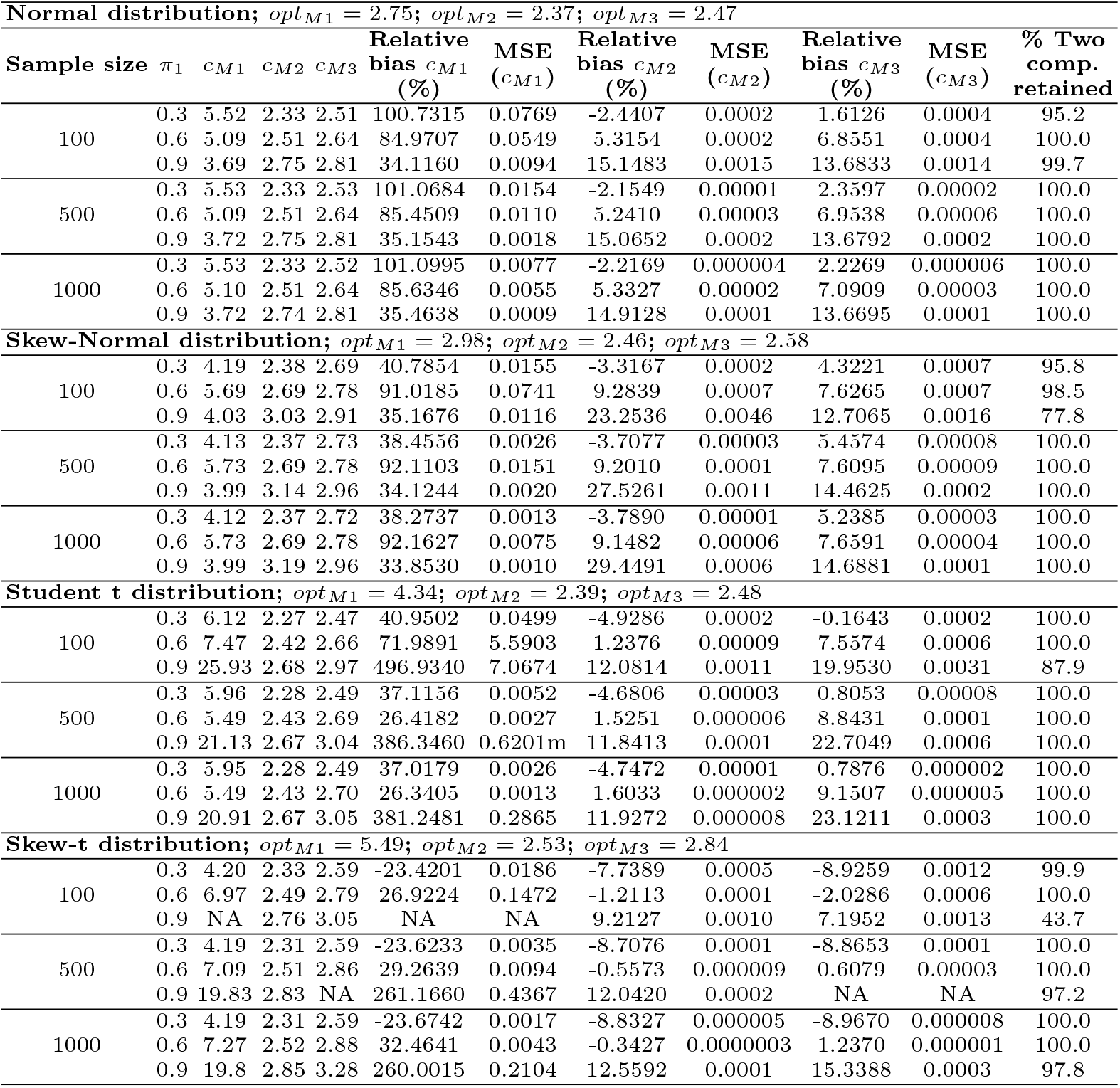
Relative bias and Mean Square Error (MSE) of the 99.9%-quantile method (M1); minimum of mixture densities method (M2) and conditional probability method (M3) for the *S*^*tri*^ antigen. *opt*_*M*1_ denotes the theoretical cutoff point for the 99.9%-quantile; *opt*_*M*2_ denotes the theoretical cutoff point for the minimum of the density mixture method; *opt*_*M*3_ denotes the theoretical cutoff point for conditional probability method. *π*_1_ denotes the weight of the seronegative population; *c*_*M*1_ denotes the cutoff estimated by M1 method after N=1000 simulations; *c*_*M*2_ denotes the cutoff estimated by M2 method after N=1000 simulations; *c*_*M*3_ denotes the cutoff estimated by M3 method after N=1000 simulations.

| Normal distribution; $opt_{M1} = 2.75$; $opt_{M2} = 2.37$; $opt_{M3} = 2.47$ | | | | | | | | | | | | |
| --- | --- | --- | --- | --- | --- | --- | --- | --- | --- | --- | --- | --- |
| Sample size | $\pi_1$ | $c_{M1}$ | $c_{M2}$ | $c_{M3}$ | Relative bias $c_{M1}$ (%) | MSE ($c_{M1}$) | Relative bias $c_{M2}$ (%) | MSE ($c_{M2}$) | Relative bias $c_{M3}$ (%) | MSE ($c_{M3}$) | % Two comp. retained | |
| 100 | 0.3 | 5.52 | 2.33 | 2.51 | 100.7315 | 0.0769 | -2.4407 | 0.0002 | 1.6126 | 0.0004 | 95.2 |  |
|  | 0.6 | 5.09 | 2.51 | 2.64 | 84.9707 | 0.0549 | 5.3154 | 0.0002 | 6.8551 | 0.0004 | 100.0 |  |
|  | 0.9 | 3.69 | 2.75 | 2.81 | 34.1160 | 0.0094 | 15.1483 | 0.0015 | 13.6833 | 0.0014 | 99.7 |  |
| 500 | 0.3 | 5.53 | 2.33 | 2.53 | 101.0684 | 0.0154 | -2.1549 | 0.00001 | 2.3597 | 0.00002 | 100.0 |  |
|  | 0.6 | 5.09 | 2.51 | 2.64 | 85.4509 | 0.0110 | 5.2410 | 0.00003 | 6.9538 | 0.00006 | 100.0 |  |
|  | 0.9 | 3.72 | 2.75 | 2.81 | 35.1543 | 0.0018 | 15.0652 | 0.0002 | 13.6792 | 0.0002 | 100.0 |  |
| 1000 | 0.3 | 5.53 | 2.33 | 2.52 | 101.0995 | 0.0077 | -2.2169 | 0.000004 | 2.2269 | 0.000006 | 100.0 |  |
|  | 0.6 | 5.10 | 2.51 | 2.64 | 85.6346 | 0.0055 | 5.3327 | 0.00002 | 7.0909 | 0.00003 | 100.0 |  |
|  | 0.9 | 3.72 | 2.74 | 2.81 | 35.4638 | 0.0009 | 14.9128 | 0.0001 | 13.6695 | 0.0001 | 100.0 |  |
| Skew-Normal distribution; $opt_{M1} = 2.98$; $opt_{M2} = 2.46$; $opt_{M3} = 2.58$ | | | | | | | | | | | | |
| 100 | 0.3 | 4.19 | 2.38 | 2.69 | 40.7854 | 0.0155 | -3.3167 | 0.0002 | 4.3221 | 0.0007 | 95.8 |  |
|  | 0.6 | 5.69 | 2.69 | 2.78 | 91.0185 | 0.0741 | 9.2839 | 0.0007 | 7.6265 | 0.0007 | 98.5 |  |
|  | 0.9 | 4.03 | 3.03 | 2.91 | 35.1676 | 0.0116 | 23.2536 | 0.0046 | 12.7065 | 0.0016 | 77.8 |  |
| 500 | 0.3 | 4.13 | 2.37 | 2.73 | 38.4556 | 0.0026 | -3.7077 | 0.00003 | 5.4574 | 0.00008 | 100.0 |  |
|  | 0.6 | 5.73 | 2.69 | 2.78 | 92.1103 | 0.0151 | 9.2010 | 0.0001 | 7.6095 | 0.00009 | 100.0 |  |
|  | 0.9 | 3.99 | 3.14 | 2.96 | 34.1244 | 0.0020 | 27.5261 | 0.0011 | 14.4625 | 0.0002 | 100.0 |  |
| 1000 | 0.3 | 4.12 | 2.37 | 2.72 | 38.2737 | 0.0013 | -3.7890 | 0.00001 | 5.2385 | 0.00003 | 100.0 |  |
|  | 0.6 | 5.73 | 2.69 | 2.78 | 92.1627 | 0.0075 | 9.1482 | 0.00006 | 7.6591 | 0.00004 | 100.0 |  |
|  | 0.9 | 3.99 | 3.19 | 2.96 | 33.8530 | 0.0010 | 29.4491 | 0.0006 | 14.6881 | 0.0001 | 100.0 |  |
| Student t distribution; $opt_{M1} = 4.34$; $opt_{M2} = 2.39$; $opt_{M3} = 2.48$ | | | | | | | | | | | | |
| 100 | 0.3 | 6.12 | 2.27 | 2.47 | 40.9502 | 0.0499 | -4.9286 | 0.0002 | -0.1643 | 0.0002 | 100.0 |  |
|  | 0.6 | 7.47 | 2.42 | 2.66 | 71.9891 | 5.5903 | 1.2376 | 0.00009 | 7.5574 | 0.0006 | 100.0 |  |
|  | 0.9 | 25.93 | 2.68 | 2.97 | 496.9340 | 7.0674 | 12.0814 | 0.0011 | 19.9530 | 0.0031 | 87.9 |  |
| 500 | 0.3 | 5.96 | 2.28 | 2.49 | 37.1156 | 0.0052 | -4.6806 | 0.00003 | 0.8053 | 0.00008 | 100.0 |  |
|  | 0.6 | 5.49 | 2.43 | 2.69 | 26.4182 | 0.0027 | 1.5251 | 0.000006 | 8.8431 | 0.0001 | 100.0 |  |
|  | 0.9 | 21.13 | 2.67 | 3.04 | 386.3460 | 0.6201m | 11.8413 | 0.0001 | 22.7049 | 0.0006 | 100.0 |  |
| 1000 | 0.3 | 5.95 | 2.28 | 2.49 | 37.0179 | 0.0026 | -4.7472 | 0.00001 | 0.7876 | 0.000002 | 100.0 |  |
|  | 0.6 | 5.49 | 2.43 | 2.70 | 26.3405 | 0.0013 | 1.6033 | 0.000002 | 9.1507 | 0.000005 | 100.0 |  |
|  | 0.9 | 20.91 | 2.67 | 3.05 | 381.2481 | 0.2865 | 11.9272 | 0.000008 | 23.1211 | 0.0003 | 100.0 |  |
| Skew-t distribution; $opt_{M1} = 5.49$; $opt_{M2} = 2.53$; $opt_{M3} = 2.84$ | | | | | | | | | | | | |
| 100 | 0.3 | 4.20 | 2.33 | 2.59 | -23.4201 | 0.0186 | -7.7389 | 0.0005 | -8.9259 | 0.0012 | 99.9 |  |
|  | 0.6 | 6.97 | 2.49 | 2.79 | 26.9224 | 0.1472 | -1.2113 | 0.0001 | -2.0286 | 0.0006 | 100.0 |  |
|  | 0.9 | NA | 2.76 | 3.05 | NA | NA | 9.2127 | 0.0010 | 7.1952 | 0.0013 | 43.7 |  |
| 500 | 0.3 | 4.19 | 2.31 | 2.59 | -23.6233 | 0.0035 | -8.7076 | 0.0001 | -8.8653 | 0.0001 | 100.0 |  |
|  | 0.6 | 7.09 | 2.51 | 2.86 | 29.2639 | 0.0094 | -0.5573 | 0.000009 | 0.6079 | 0.00003 | 100.0 |  |
|  | 0.9 | 19.83 | 2.83 | NA | 261.1660 | 0.4367 | 12.0420 | 0.0002 | NA | NA | 97.2 |  |
| 1000 | 0.3 | 4.19 | 2.31 | 2.59 | -23.6742 | 0.0017 | -8.8327 | 0.000005 | -8.9670 | 0.000008 | 100.0 |  |
|  | 0.6 | 7.27 | 2.52 | 2.88 | 32.4641 | 0.0043 | -0.3427 | 0.0000003 | 1.2370 | 0.000001 | 100.0 |  |
|  | 0.9 | 19.8 | 2.85 | 3.28 | 260.0015 | 0.2104 | 12.5592 | 0.0001 | 15.3388 | 0.0003 | 97.8 |  |

In general it is found that as the sample size increases, both the relative error and the root mean square error tend to decrease. It is also found that for small samples and extreme *π*_1_ values (*π*_1_ = 0.3 or *π*_1_ = 0.9), the models tend to have some difficulty in identifying a seronegative and seropositive population. This is a result that alerts to the existence of possible false positives and false negatives in the case of small samples.

In situations where there is an ongoing vaccination plan and therefore the majority of the population is seropositive (e.g. *π*_1_ = 0.9) it is important to know if it is possible to identify seronegative individuals in this population given the time of immunization. If the timing of immunization is short, it is important to identify these individuals early in order to take action and prevent a further increase in infections.

## 5 Conclusions

The purpose of this study was to use a flexible class of mixture models to antibody data against the SARS-CoV-2 virus. In particular, we used a class of models that allows capturing the skewness present in this type of data, namely the Skew-Normal and Skew-t distributions.

It has become clear that diagnostic tests play a key role in the early identification of infected individuals, allowing us to act to control a pandemic by isolating and tracing the contacts of an infected person. Diagnostic tests can classify an individual as seronegative or seropositive by defining a cutoff point that can take on different values depending on the technique used by the manufacturer to develop the test. Most of the time, this cutoff point is relaxed and is calculated using the 3*σ*-rule, which assumes that the underlying distribution of the data is Normal. However, as we have seen in our application, this assumption cannot always be made, making this method unfeasible.

Note that this study has the advantage that the true cases and controls of the infection are known, allowing us to compare different methods for obtaining the cutoff point that allows classifying an individual as seropositive.

In [9], three methods for obtaining the cutoff point had been presented that could not yet be validated because the true infection status of the individuals was not known. In this sense, we proceeded to use these methods in this study, and it was verified that the three methods under analysis present high accuracy, compared to methods used in literature, namely through the empirical ROC curve. However, the proposed methods proved to be more specific than sensitive. Note that the performance of the method based on the 99.9% probability quantile may be overestimated, especially when the fitted distribution corresponds to a heavy-tailed distribution (such as the Skew-t distribution). This is because the calculation of this quantile involves only and exclusively the population of seropositive individuals, so that if the distribution is too skewed to the right, then the seropositive population is totally absorbed by this quantile.

When a new virus is present in the population, there is a natural tendency for the proportion of susceptible individuals to be much higher than the seropositive individuals. This is the phase in which early identification of the infected people is essential for pandemic control, although total control of the spread of the virus only occurs when there is vaccination or eradication of the virus. In this sense, with the simulation study developed in this work, we intend to analyze the pandemic evolution scenarios and understand the behavior of different methods for determining the cutoff point. It was found that as the sample size increases, there is a tendency for the relative error and the mean square error of the cutoff point estimates in skewed distributions to decrease, while this tendency is not linear in the case of the usual symmetric distributions (Normal and Student t). This fact may be due to the fact that symmetrical distributions are not the most appropriate for these types of data, or even that the proposed methods should not be used when considering the usual distributions.

As we expected, for small sample sizes and for large imbalances in the serological populations, the proposed models were found to have problems in identifying two components. Note that in the case of skewed distributions, it will be natural that if the weight of the seronegative population is very high, then observations relating to the seropositive population are considered false negatives and false positives otherwise.

A limitation of this study is the fact that the adjustment of the different mixture models was performed using the same distribution for the two components (through the package mixsmsn). If the components of the mixture model were distinct, this would have a direct implication on the estimated cutoff points. However, the package that would allow this analysis is now discontinued.

In conclusion, we recommend the use of mixture models based on distributions of the SMSN family for the analysis of serological data given the flexibility of these models, as well as the use of the proposed methods for determining cutoff points as an alternative to the method based on the 3*σ* rule.

## Data Availability

All data produced are available online at https://github.com/MWhite-InstitutPasteur/SARSCoV2SeroDXphase2

